# Adjusting COVID-19 seroprevalence survey results to account for test sensitivity and specificity

**DOI:** 10.1101/2021.03.04.21252939

**Authors:** Mark J. Meyer, Shuting Yan, Samantha Schlageter, John D. Kraemer, Eli S. Rosenberg, Michael A. Stoto

## Abstract

Population-based seroprevalence surveys can provide useful estimates of the number of individuals previously infected with SARS-CoV-2 and still susceptible as well as contribute to better estimates of the case fatality rate and other measures of COVID-19 severity. No serological test is 100% accurate, however, and the standard correction that epidemiologists use to adjust estimates relies on estimates of the test sensitivity and specificity often based on small validation studies. This paper develops a fully Bayesian approach to adjust observed prevalence estimates for sensitivity and specificity. Application to a seroprevalence survey conducted in New York State in 2020 demonstrates that this approach results in more realistic – and narrower – credible interval than the standard sensitivity analysis using confidence interval endpoints. In addition, the model permits incorporating data on the geographical distribution of reported case counts to create informative priors on the cumulative incidence to produce estimates and credible intervals for smaller geographic areas than often can be precisely estimated with seroprevalence surveys.

## Introduction

Tracking the spread of COVID-19 through communities and identifying the virus’ epidemiological characteristics requires a variety of surveillance systems. Most prominent among these are the daily counts of cases and deaths that are reported in the media and used as metrics to guide decisions such as re-opening schools. However, because these counts depend on the demand for and availability of testing as well as other variable factors, they are misleading representations of trends in the incidence of cases and deaths (1–3).

To complement these data, some communities undertake seroprevalence surveys, in which a representative sample of individuals from a defined population are tested to identify the presence of antibodies indicating a previous infection with SARS-CoV2, the virus that causes COVID-19 (4–6). Seroprevalence surveys can be conducted in geographically-defined populations (e.g. New York State (7), New York City (8), England (9), or Spain (10), people receiving medical care at a particular site (e.g. women delivering babies at specific clinics in New York City (11) or undergoing dialysis nationally (12)), blood samples collected for routine screening (13,14), or as part of ongoing surveillance at nursing homes and other high likelihood of exposure sites.

When infection reliably produces an antibody response and case-fatality rates are relatively small, seroprevalence approximates cumulative incidence over the average period of detectable antibody. Because SARS-CoV-2 infection likely produces detectable antibodies for months after the infection has cleared, seroprevalence enables a reasonable estimate of recent cumulative infections. Further, seroprevalence surveys are used to estimate the number of persons still susceptible and progress towards herd immunity, a use for which mortality can be neglected. Finally, if reasonable estimates of mortality are available—and mortality is often easier to measure through routine surveillance than incidence—seroprevalence surveys can also contribute to better estimates of the case fatality rate and other measures of disease severity, to get a sense of how close a population is to achieving herd immunity, which are critical for parameterizing simulation models and informing policies (15–18).

Conducting and analyzing a seroprevalence study, however, can be challenging. The most prominent problem, of course, is identifying a representative sample, either by random sampling or other means, and ensuring that individuals who have been infected are neither more or less likely to be included. Second, when prevalence is low, which to date is typically the case with COVID-19, the number of positive tests in a sample is small. This is exacerbated, when the sample is broken down by subgroups that correspond to, for example, geographic areas within a state—leading to substantial uncertainty about estimates.

Beyond that, even the best tests are not 100% accurate; false negatives and false positives are to be expected (19). Epidemiologists address this with a standard correction formula based on Bayes’ Rule and estimates of the test sensitivity and specificity:

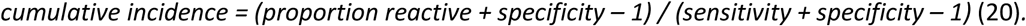

The estimates of sensitivity and specificity, however, are often based on small validation studies. Rosenberg and colleagues, for instance, estimated the specificity of their assay (the proportion of non-cases that test negative) as 99.75% with a 95% confidence interval of 96.1% to 100%. The uncertainty about the sensitivity of their assay (the proportion of actual cases that test positive) was much larger, with a base estimate of 87.9% and a 95% confidence interval of 83.7% to 92.1%. State-wide (and adjusting for sampling and demographic factors), Rosenberg and colleagues estimated a cumulative incidence of 14.0% (95% CI: 13.3-14.7%) using the mean sensitivity and specificity. In sensitivity analyses at the extremes of test characteristics, however, cumulative incidence ranged broadly from 9.8% to 15.0% (7). There is an additional problem with the standard correction formula: when the actual prevalence is low compared to 1-specificity, a negative number can result. For example, if the proportion reactive was 2%, the extreme values of the Rosenberg and colleagues test characteristics would yield a cumulative incidence range from −0.0215 to 0.0239. Since cumulative incidence cannot be negative, and because using the extreme values of the 95% confidence intervals for both sensitivity and specificity results does not yield a 95% confidence interval for the adjustment, the resulting intervals cannot be described in probabilistic terms.

The primary objective of this paper is to develop a fully Bayesian approach to adjust observed prevalence estimates for sensitivity and specificity with a more realistic – and narrower – credible interval than the standard sensitivity analysis using confidence interval endpoints. In addition, the model we have developed permits incorporating data on the geographical distribution of reported case counts to create informative priors on the cumulative incidence to produce estimates and credible intervals for smaller geographic areas than often can be precisely estimated with seroprevalence surveys.

## Methods

### Data

This study re-analyzes seroprevalence data produced by Rosenberg and colleagues (7). The seroprevalence data were collected between April 19 and 28, 2020, and have been fully described previously (7). Consistent with this analysis, data for New York State are categorized by county, region, and super-region. In Figure 1, the 62 counties are outlined in white, the 10 regions are outlined in grey, and the 4 super-regions are outlined in black. The regions include the Capital Region, Central New York, Finger Lakes, Hudson Valley, Long Island, Mohawk Valley, New York City, North Country, Southern Tier, and Western New York (21). For the purpose of this study, Westchester and Rockland Counties were separated from the Hudson Valley Region and were treated as their own region. The super-regions, which were defined to reflect differences in COVID-19 epidemiology, include New York City, Westchester and Rockland Counties plus Long Island, and the remainder of the state (“Rest of State”).

As a basis for the prior distributions, we calculated the cumulative reported cases for the region by grouping the cumulative reported cases for each county in New York State on April 11, 2020. The literature estimates a mean of 4 days for symptom onset after infection (22), and a mean of 9 days for diagnosis after symptom onset (or 13 days for diagnosis after infection) during the early stages of the epidemic in New York (23). Given that Rosenburg and colleagues estimated seroprevalence results that equate to cumulative incidence through approximately March 29, 2020, we analyzed cumulative reported cases for New York State on April 11, 2020 which is 13 days after estimated infection and 9 days after estimated diagnosis.

**Figure 1.**
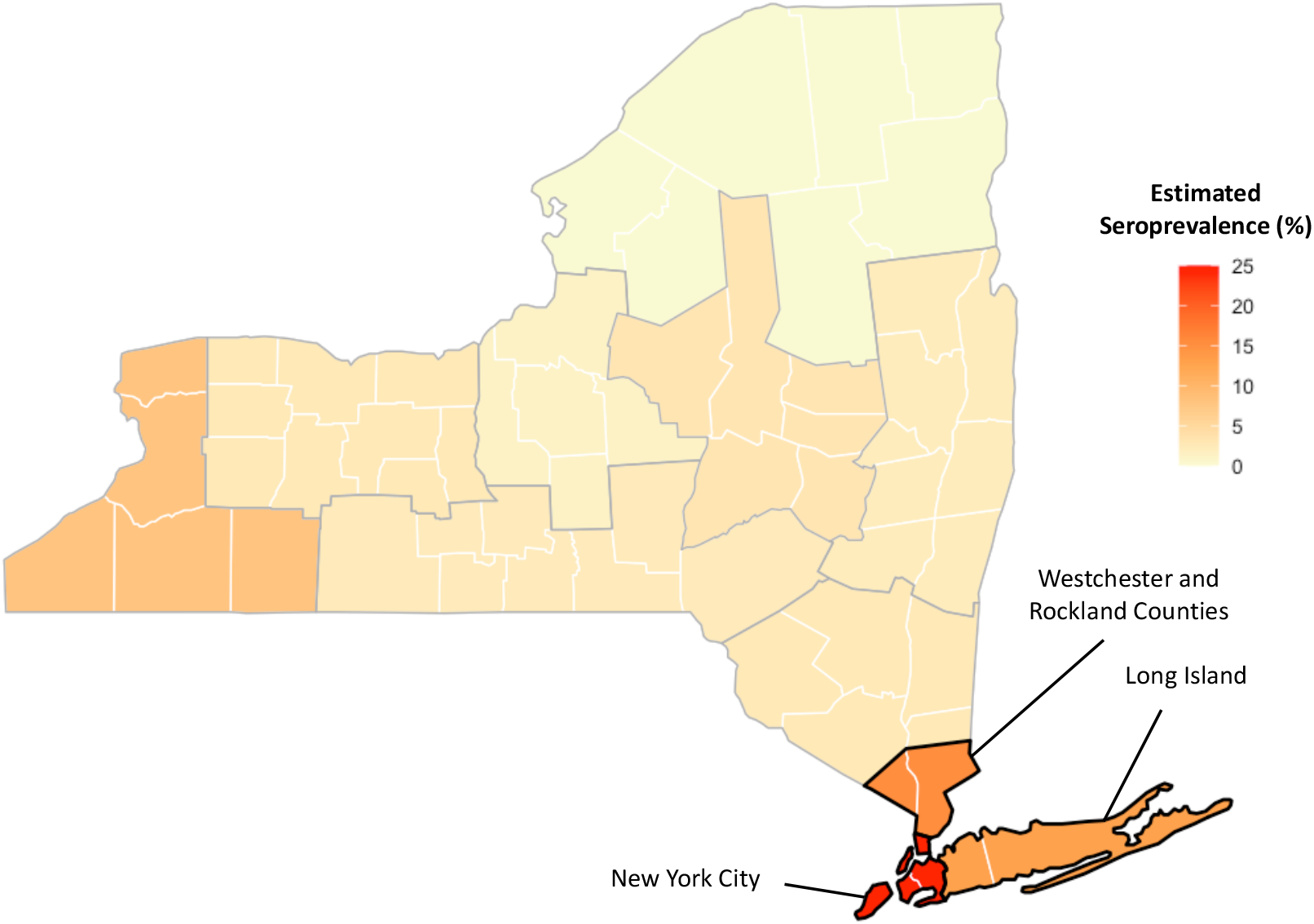
Estimated Seroprevalence by Region.

To calculate the cumulative reported cases per 10,000 for each region, the cumulative reported cases on April 11, 2020 for the counties were grouped by region. The 2019 annual population estimates for the counties were also grouped by distinct region (24). The cumulative reported cases for each county were summed by region and divided by the region’s estimated population, creating a cumulative case rate for each region as represented in Table 1 (25). The county cumulative reported cases on April 11, 2020 were also grouped by super-region using the same approach. These estimates were then multiplied by 10 to account for estimated under-ascertainment of reported cases based on previous estimates of the spring epidemic (12,13,26). These estimates then formed the basis of the prior estimate for true seroprevalence in each jurisdiction as described below.

**Table 1.** Cumulative reported cases per 10,000 by region and super region as of April 11, 2020.

| Panel a. By region | Cases per 10,000 |
| --- | --- |
| Capital Region | 9.23 |
| Central New York | 7.26 |
| Finger Lakes | 8.62 |
| Westchester & Rockland Counties | 202.63 |
| Hudson Valley (*excluding Westchester & Rockland Counties*) | 77.7 |
| Long Island | 149.87 |
| Mohawk Valley | 10.45 |
| New York City | 117.92 |
| North Country | 4.79 |
| Southern Tier | 8.51 |
| Western New York | 12.38 |

| Panel b. By super-region | Cases per 10,000 |
| --- | --- |
| New York City | 117.92 |
| Westchester & Rockland Counties | 202.63 |
| Long Island | 149.87 |
| Rest of State | 19.3 |

### Statistical Analysis

Our primary inferential goal is the true seroprevalence, that is the proportion of individuals who have detectable antibodies in each region and super region. To obtain this, we let *Ts*_*ij*_ denote the true seroprevalence in the *i* th sub-region of the *j*th super-region, *i* = 1, …,*m*_*j*_ and *j* = 1, 2, 3. Super-regions are as defined in Figure 1. Also define the observed seroprevalence in the *ij*th region to be 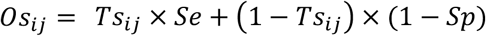 for the test sensitivity, *Se*, and specificity, *Sp*. The likelihood, across all regions, is

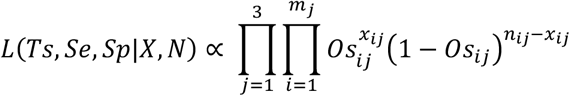

for *x* positive seroprevalence tests out of *n* total tests performed in each *ij* region. We place the following priors on the sensitivity and specificity:

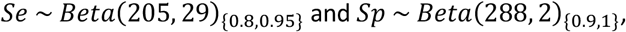

where {} denotes truncation to the specified regions. We chose the parameters of these Beta distributions so that the means of these distributions are the same as the sensitivity and specificity values used by Rosenberg and colleagues and the variances represented a reasonable range of values for these test characteristics (7). Truncated distributions are re-weighted to integrate to 1, thus these priors are proper.

Geographic regions may have similar seroprevalence rates to their neighbors due to similar socio-demographic and geographic factors. Hierarchical priors across the super-regions accommodate this structure. Weakly informative and informative priors incorporate information from regional-specific cumulative case counts found in Table 1. Thus, for each region within a super-region, the true seroprevalences are assumed to be independent and identically distributed samples from the super-region specific prior. We use a weakly informative prior for our primary analysis but also employ both a non-informative and more informative prior in sensitivity analyses.

For hierarchical priors based on the geographical clusters defined by the super-regions, *j* = 1 denotes New York City, *j* = 2 denotes Rockland and Westchester counties along with Long Island, and *j* = 3 denotes the rest of the state. The hierarchical priors are then

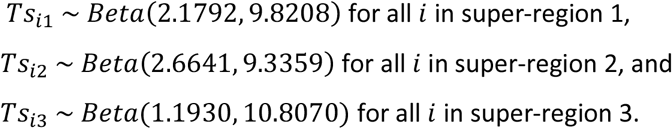

We chose the parameters of these Beta distributions so that the mean was the estimated prevalence based on reported cases in each super-region. These priors are considered weakly informative because despite incorporating information on the location of the parameter, the variance of each prior is relatively large. For the uniform prior seroprevalence and informative prior seroprevalence models, please see the Methods Appendix. Graphical depictions of all priors are in the Supplementary Material.

Using this model, we first calculate seroprevalence estimates for each region. We then construct super-region and statewide estimates as averages of the region-specific posterior seroprevalence estimates weighted by the proportion of the super-region or state’s population that lives in each region. Data for the population-based weights comes from the U.S. Census Bureau 2019 Population Estimates, provided by the New York State Department of Labor (25).

Model estimates are sampled using the NIMBLE package in R (27,28), which performs Markov Chain Monte Carlo (MCMC) simulation. Sample code to implement the model is available in the appendix while we make our full code freely available online at https://github.com/markjmeyer/BPA. For all models under consideration, we sample four separate chains each of length 200,000, retaining 100,000 samples for conducting inference. We monitor convergence using the Gelman-Rubin diagnostic (29) and traceplots. All parameters were judged to have properly converged. Traceplots and tables of convergence diagnostics are in the Supplementary Material.

## Results

Prior estimates for true seroprevalence in each super-region, sensitivity, and specificity are shown Figure 1 and supplemental table 1. Region-specific posterior estimates of seroprevalence, adjusted for sensitivity and specificity, are shown Table 2. True seroprevalence estimates range from 1.0% (95% credible interval [CrI] 0.1-2.9%) to 25.0% (95% CrI 23.3-26.9%). True seroprevalence was estimated at 15.3% (95% CrI 12.7-18.1%) in Westchester and Rockland Counties, 12.9% (95% CrI 11.1-14.7%) in Long Island, and 7.9% (95% CrI 6.2-9.9%) in Western New York. True seroprevalence is estimated below 5% as of the date of data collection in all other regions. The weighted estimate of the true seroprevalence in New York state is 14.8% (95% CrI 13.7-16.0%). The weighted estimate of the true seroprevalence in the Westchester County, Rockland County, and Long Island super-region was 13.6% (95% CrI 12.1-15.3%); in the super-region consisting of the remainder of the state (not including New York City), estimated true seroprevalence is 3.4% (95% CrI 2.5-4.2%).

**Table 2:**
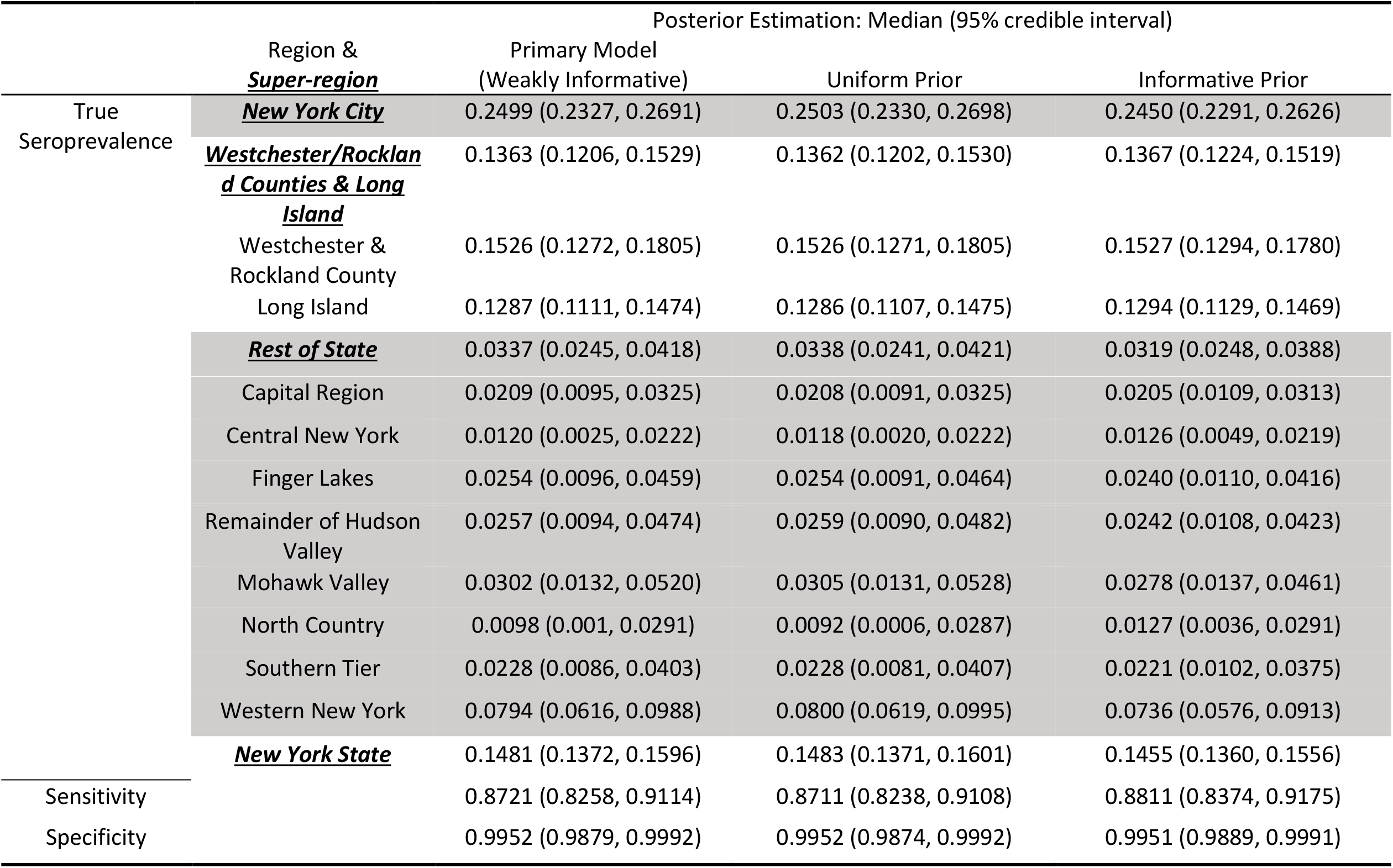
Posterior Estimates of the true prevalence on March 29, 2020 for each region, sensitivity, and specificity under all models.

|  | Region &<br>***Super-region*** | Posterior Estimation: Median (95% credible interval) |  |  |
| --- | --- | --- | --- | --- |
|  |  | Primary Model<br>(Weakly Informative) | Uniform Prior | Informative Prior |
| True<br>Seroprevalence | **<u>New York City</u>** | 0.2499 (0.2327, 0.2691) | 0.2503 (0.2330, 0.2698) | 0.2450 (0.2291, 0.2626) |
|  | **<u>Westchester/Rockland Counties & Long Island</u>** | 0.1363 (0.1206, 0.1529) | 0.1362 (0.1202, 0.1530) | 0.1367 (0.1224, 0.1519) |
|  | Westchester &<br>Rockland County | 0.1526 (0.1272, 0.1805) | 0.1526 (0.1271, 0.1805) | 0.1527 (0.1294, 0.1780) |
|  | Long Island | 0.1287 (0.1111, 0.1474) | 0.1286 (0.1107, 0.1475) | 0.1294 (0.1129, 0.1469) |
|  | **<u>Rest of State</u>** | 0.0337 (0.0245, 0.0418) | 0.0338 (0.0241, 0.0421) | 0.0319 (0.0248, 0.0388) |
|  | Capital Region | 0.0209 (0.0095, 0.0325) | 0.0208 (0.0091, 0.0325) | 0.0205 (0.0109, 0.0313) |
|  | Central New York | 0.0120 (0.0025, 0.0222) | 0.0118 (0.0020, 0.0222) | 0.0126 (0.0049, 0.0219) |
|  | Finger Lakes | 0.0254 (0.0096, 0.0459) | 0.0254 (0.0091, 0.0464) | 0.0240 (0.0110, 0.0416) |
|  | Remainder of Hudson<br>Valley | 0.0257 (0.0094, 0.0474) | 0.0259 (0.0090, 0.0482) | 0.0242 (0.0108, 0.0423) |
|  | Mohawk Valley | 0.0302 (0.0132, 0.0520) | 0.0305 (0.0131, 0.0528) | 0.0278 (0.0137, 0.0461) |
|  | North Country | 0.0098 (0.001, 0.0291) | 0.0092 (0.0006, 0.0287) | 0.0127 (0.0036, 0.0291) |
|  | Southern Tier | 0.0228 (0.0086, 0.0403) | 0.0228 (0.0081, 0.0407) | 0.0221 (0.0102, 0.0375) |
|  | Western New York | 0.0794 (0.0616, 0.0988) | 0.0800 (0.0619, 0.0995) | 0.0736 (0.0576, 0.0913) |
|  | **<u>New York State</u>** | 0.1481 (0.1372, 0.1596) | 0.1483 (0.1371, 0.1601) | 0.1455 (0.1360, 0.1556) |
| Sensitivity |  | 0.8721 (0.8258, 0.9114) | 0.8711 (0.8238, 0.9108) | 0.8811 (0.8374, 0.9175) |
| Specificity |  | 0.9952 (0.9879, 0.9992) | 0.9952 (0.9874, 0.9992) | 0.9951 (0.9889, 0.9991) |

Credible intervals are consistently narrower when estimated using a fully Bayesian approach than when employing the customary correction for estimated sensitivity and specificity to confidence interval endpoints around seroprevalence estimates (Table 3). For example, when the unadjusted seroprevalence is 20%, as in New York City, the interval width is 3.6 percentage points using the fully Bayesian approach compared to 5.6 percentage points with the customary approach. When the unadjusted seroprevalence is 12%, as in the Rockland, Westchester and Long Island super-region, the results are similar. When the unadjusted seroprevalence is low, for example 2%, a typical value in most regions of the state, the results are different. There is a similar relative reduction in the width of the interval, from 4.5 to 2.3 percentage points. More importantly, the sensitivity and specificity-corrected lower bound estimate is 1.0% using the fully Bayesian approach, as opposed to a negative number, - 2.1%, using the customary approach.

**Table 3.** Comparison of 95% credible intervals based on the fully Bayesian approach (primary model) and a sensitivity analysis based on the customary correction for sensitivity and specificity

|  | Mean & 95% C.I. |  |  |
| --- | --- | --- | --- |
| *Observed antibody + = 2%* |  |  |  |
| Sensitivity | 92.10% | 87.90% | 83.70% |
| Specificity | 96.10% | 99.75% | 100% |
| Sensitivity analysis | **-0.0215** | **0.0200** | **0.0239** |
| 95% credible interval | 0.0096 | 0.0209 | 0.0324 |
| *Observed antibody + = 12%* |  |  |  |
| Sensitivity analysis | **0.0918** | **0.1341** | **0.1434** |
| 95% credible interval | 0.1110 | 0.1287 | 0.1473 |
| *Observed antibody + = 20%* |  |  |  |
| Sensitivity analysis | **0.1825** | **0.2253** | **0.2389** |
| 95% credible interval | 0.2326 | 0.2498 | 0.2690 |

Estimates of true seroprevalence are robust to the choice of prior distributions (Table 2). The maximum deviation in true seroprevalence estimates between the primary model with a weakly informative prior and the sensitivity analysis using uniform priors is 0.06 percentage points in Western New York. Deviation was slightly greater when the primary model was compared to the sensitivity analysis using informative priors, with Western New York again having the largest deviation (0.6 percentage points). Credible interval widths are slightly wider with estimates using uniform priors and slightly narrower with estimates using informative priors but differences are not meaningful.

Figure 2 illustrates the effect using hierarchical weakly informative and non-informative priors for the entire “Rest of State” super-region. The customary correction has the effect of adjusting the proportion reactive up if it is greater than 2.0% and down otherwise. In general, the Bayesian model moves the estimates back towards the mode of the super-region’s hierarchical prior. The non-informative prior for our model forms the basis of comparison as its mode is undefined. For the weakly informative prior, the “Rest of State” prior mode is 1.9% and model estimates shift toward this value when compared to those under the non-informative prior. This is the case for all super-regions, but because this region has lower seroprevalence, the influence of the prior is more noticeable. It is most apparent for Western New York, which has a higher seroprevalence than the rest of the super-region, and for North Country, which has the smallest sample (299 test) and number reactive (3). The “Rest of State” super-region may be too large resulting in a hierarchical prior that smooths over sub-regional differences. However, based on the available information, the weakly informative prior represents our “best guess” for this region. Ultimately, the data from regions like Western New York overwhelm the information imparted by prior providing a better basis for future Bayesian analyses of seroprevalence data in New York.

**Figure 2.**
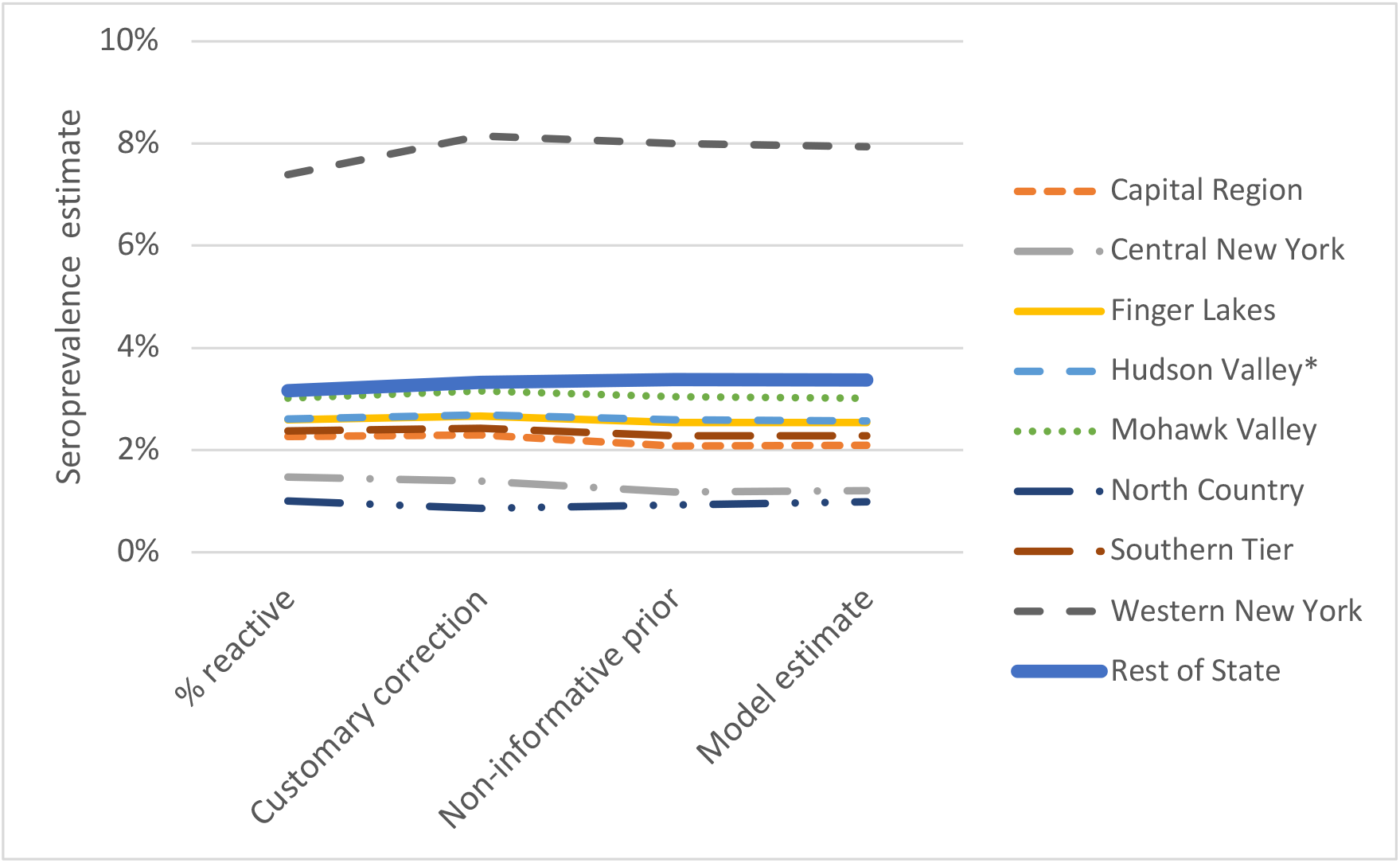
Seroprevalence estimated by the proportion reactive, the customary correction, and the fully Bayesian approach with both weakly informative and non-informative priors for the regions in the “Rest of State” super-region.

## Discussion

These results demonstrate that the fully Bayesian model achieves the two goals we set out to meet. First, as shown in Table 3, the credible intervals for seroprevalence that result are substantially narrower than the intervals one would calculate using the customary correction for estimated sensitivity and specificity, and do not yield negative numbers for proportions that must be greater than zero. The customary approach, which used the extreme ends of the confidence intervals for estimated sensitivity and specificity instead of the variance for an adjusted estimator, is not a 95% confidence interval and should not be interpreted as such. The 95% credible intervals our method calculates, on the other hand, have a proper probabilistic interpretation: 95% of the posterior distribution is between the lower and upper bounds. Confidence intervals are often misinterpreted in this fashion, giving credible intervals the advantage of having a more natural and less misleading interpretation.

When it comes to accounting for sensitivity and specificity of tests used in seroprevalence surveys, actual practice varies substantially. Some studies (30) report point estimates of sensitivity and specificity, but do not say how (or whether) these estimates were used to adjust the results. Stadlbauer and colleagues report point estimates and confidence intervals based on small samples, but do not indicate how (or whether) this uncertainty in the seroprevalence estimates (8). Angulo and colleagues found that the ratio of survey-based estimates of the cumulative incidence of COVID-19 to reported cases in five states varied from 2.1 to 10.5 (31). Perhaps some of this variability is due to differences in how the authors accounted for test performance.

Havers (13) and Bajema (14) use bootstrap approaches to obtain confidence intervals. Each uses a different version of a two-stage approach where each bootstrap replicate incorporates an adjustment for sensitivity and specificity at the first stage. Havers and colleagues (13) estimate variability in sensitivity and specificity by resampling from test validation data and at the second step seroprevalence sampling variability is estimated by resampling observations from the seroprevalence survey. Their first stage substitutes the empirical, discrete distribution from their observations for the continuous distribution a Bayesian approach can employ. Bajema and colleagues (14) specify a binomial distribution based on laboratory-provided sensitivity and specificity values, sample from it, calculate false positive and negative rates, and then randomly flip positive and negative observations in each bootstrap replicate. These frameworks are partially Bayesian in nature as they seek to account for the variability in the sensitivity and specificity but do not directly model it as a fully Bayesian approach does. Fully Bayesian models can incorporate prior information about prevalence in ways that frequentist approaches cannot, and Bayesian approaches often allow complex estimates to be calculated more tractably at lower computational time.

Second, the incorporation of informative priors based on reported COVID-19 cases seems to have reasonably stabilized the estimates, especially for the low-prevalence areas of the state. The Bayesian estimates for New York City, with the highest seroprevalence rates, are barely different than the customary correction; the proportion seropositive, adjusted for sensitivity and specificity, and the Bayesian estimate are both 25.0%. For the low prevalence regions in “Rest of State,” however, adjusted seropositive proportions and the Bayesian estimates differ. In Central New York, the lowest prevalence region, they are 1.5% and 1.2% respectively; in Western New York, the corresponding estimates are 7.4% and 7.9%.

Analysts have often used simple approaches such as that described by Rothman and colleagues (20) because the formal Bayesian approach we describe have been challenging to compute. Recent advances in Bayesian software packages have now made this type of modeling more accessible to the non-technical user. To contribute to this accessibility, we made sample code available in the Supplementary Material and our full code available online.

When seroprevalence is used as a proxy for cumulative incidence, researchers need to be cognizant of the duration over which antibodies are detectible (32). Because this analysis uses data from the opening months of the COVID-19 epidemic, this is not an issue for our paper. It could be an issue for other studies done later in the epidemic. One might combine other sources of information about the trajectory of an epidemic—for example, data about hospitalizations or deaths—with evidence about the temporal dynamics of seroreversion to determine what proportion of total cases are likely to still have detectible antibody at the point seroprevalence sampling was conducted. The seroprevalence estimate would then be upweighted inverse to the estimated proportion of total cases that continue to have detectible antibody. More sophisticated approaches could incorporate separate evidence about epidemic trajectories stratified by demographic characteristics or geography. An alternative approach would be to conduct serial seroprevalence surveys. Based on the time between samples, one could estimate and account for the proportion of those previously positive who would no longer be expected to be positive in the subsequent sample. Accounting for seroreversion is a topic that would benefit from additional research.

Our approach is not limited to seroprevalence studies. Rather, it could be used equally well with prevalence surveys that seek to identify current prevalence via PCR or antigen testing. In fact, because current prevalence of SARS-CoV-2 infection is expected to be low in most populations, our approach may perform better than the standard adjustment for sensitivity and specificity because our approach produces intervals that do not extend below zero. Further, the approach could be extended to serial surveys for either active infection or seroprevalence. In this instance, estimates from earlier surveys— potentially combined with routine surveillance case counts, hospitalizations, or mortality estimates— would form the basis of informative priors for subsequent surveys.

This analysis ignored the poststratification weighting used by Rosenberg and colleagues (7) to make the non-probability sampling approach that was available during the early COVID-19 epidemic more representative of the New York population. However, the method we use could be applied to data from complex sampling designs or from non-probability sampling with weight adjustments to mitigate sampling bias. Our code requires raw seroprevalence data to be input as n/N, where n represents the number testing positive and N is the total sample size in a location. The easiest adjustment for weighted and/or clustered data would be to adjust N by dividing it by the design effect readily output by most statistical software. Then n could be adjusted so that the adjusted n/N equals the weighted seroprevalence estimate.

## Data Availability

All data for replicating the paper are provided in data tables. Statistical code is available at https://github.com/markjmeyer/BPA.

## Supplementary material

**Table 1:** Summary statistics of prior distributions on true prevalence, sensitivity, and specificity.

|  | Super-Region | Primary Model Priors |  |  | Uniform Prior Prevalence Model |  |  | Informative Prior Prevalence Model |  |  |
| --- | --- | --- | --- | --- | --- | --- | --- | --- | --- | --- |
|  |  | Mean | Variance | Standard Deviation | Mean | Variance | Standard Deviation | Mean | Variance | Standard Deviation |
| True Prevalence | Westchester & Rockland Counties, Long Island | 0.2220 | 0.0133 | 0.1153 | 0.5000 | 0.0833 | 0.2887 | 0.1730 | 0.0014 | 0.0373 |
|  | New York City | 0.1816 | 0.0114 | 0.1069 | 0.5000 | 0.0833 | 0.2887 | 0.1254 | 0.0011 | 0.0326 |
|  | Rest of State | 0.0994 | 0.0069 | 0.0830 | 0.5000 | 0.0833 | 0.2887 | 0.0287 | 0.0003 | 0.0165 |
| Sensitivity |  | 0.8761 | 0.0004 | 0.0199 | 0.8761 | 0.0004 | 0.0199 | 0.8761 | 0.0004 | 0.0199 |
| Specificity |  | 0.9931 | 0.0000 | 0.0049 | 0.9931 | 0.0000 | 0.0049 | 0.9931 | 0.0000 | 0.0049 |

**Table 2:**
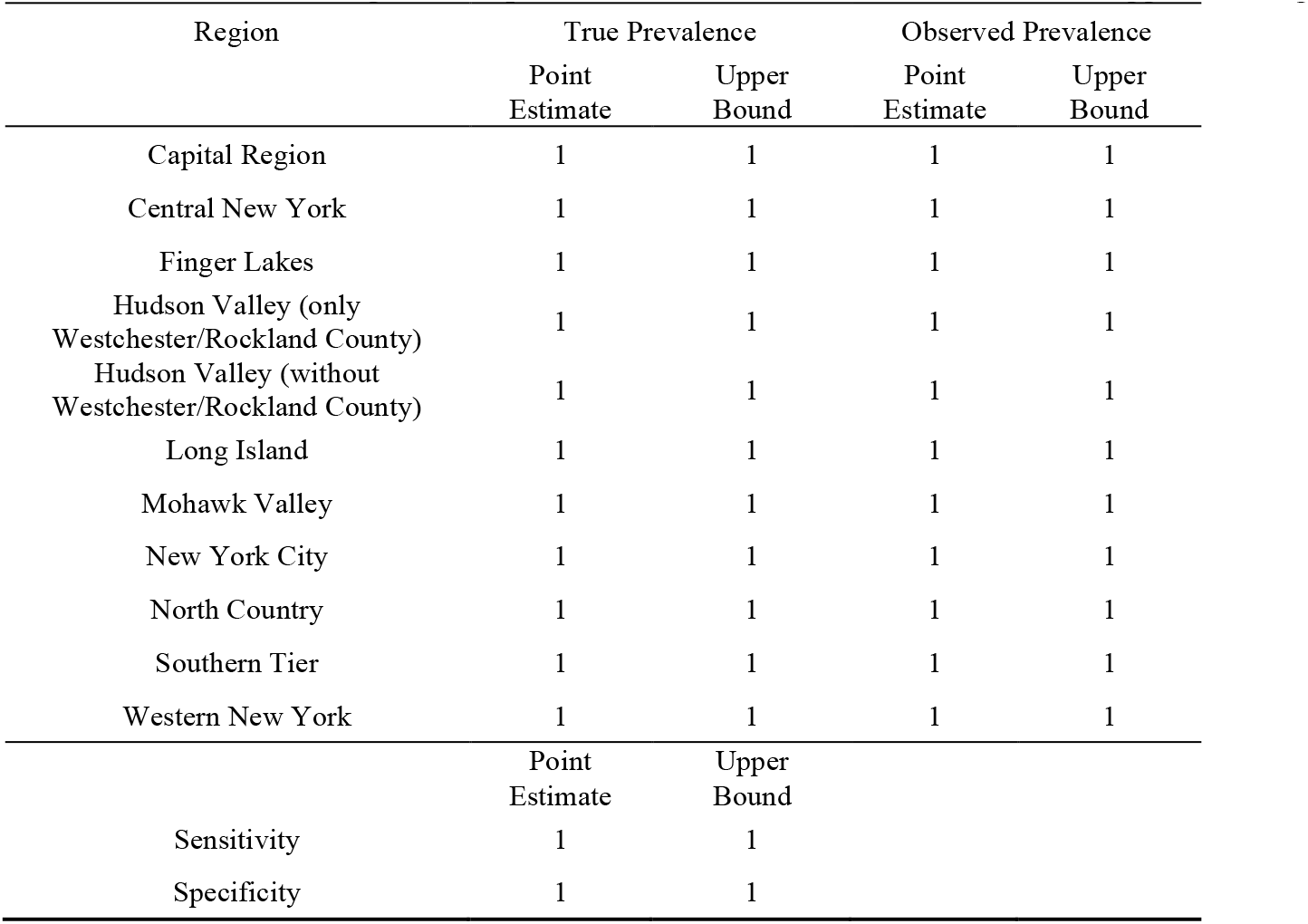
Convergence diagnostics for the primary model (weakly informative prior). The table contains point estimates and upper bound of the 95% credible interval for the Gelman-Rubin Diagnostic or potential scale reduction factor. Values of 1 suggest convergence.

| Region | True Prevalence |  | Observed Prevalence |  |
| --- | --- | --- | --- | --- |
|  | Point Estimate | Upper Bound | Point Estimate | Upper Bound |
| Capital Region | 1 | 1 | 1 | 1 |
| Central New York | 1 | 1 | 1 | 1 |
| Finger Lakes | 1 | 1 | 1 | 1 |
| Hudson Valley (only Westchester/Rockland County) | 1 | 1 | 1 | 1 |
| Hudson Valley (without Westchester/Rockland County) | 1 | 1 | 1 | 1 |
| Long Island | 1 | 1 | 1 | 1 |
| Mohawk Valley | 1 | 1 | 1 | 1 |
| New York City | 1 | 1 | 1 | 1 |
| North Country | 1 | 1 | 1 | 1 |
| Southern Tier | 1 | 1 | 1 | 1 |
| Western New York | 1 | 1 | 1 | 1 |
|  | Point Estimate | Upper Bound |  |  |
| Sensitivity | 1 | 1 |  |  |
| Specificity | 1 | 1 |  |  |

**Table 3:**
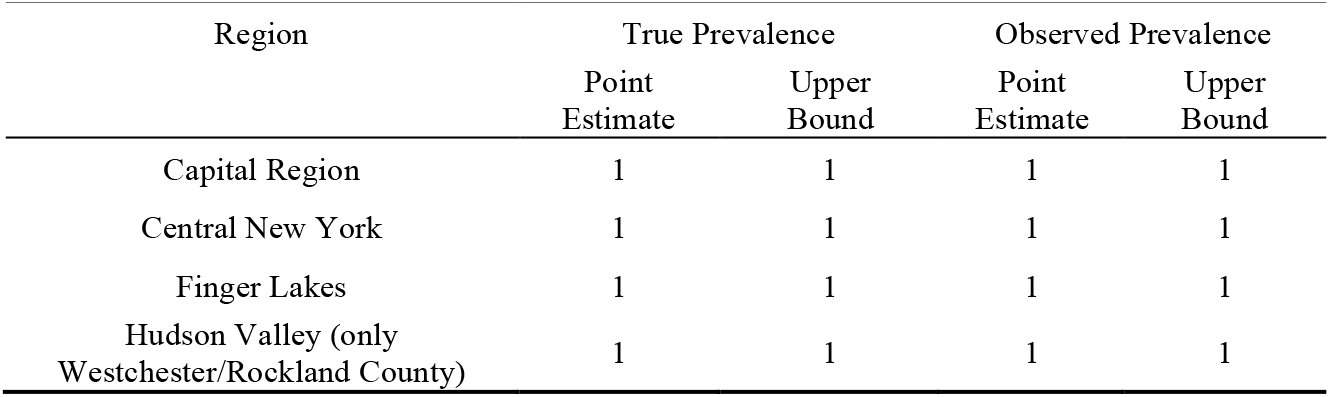
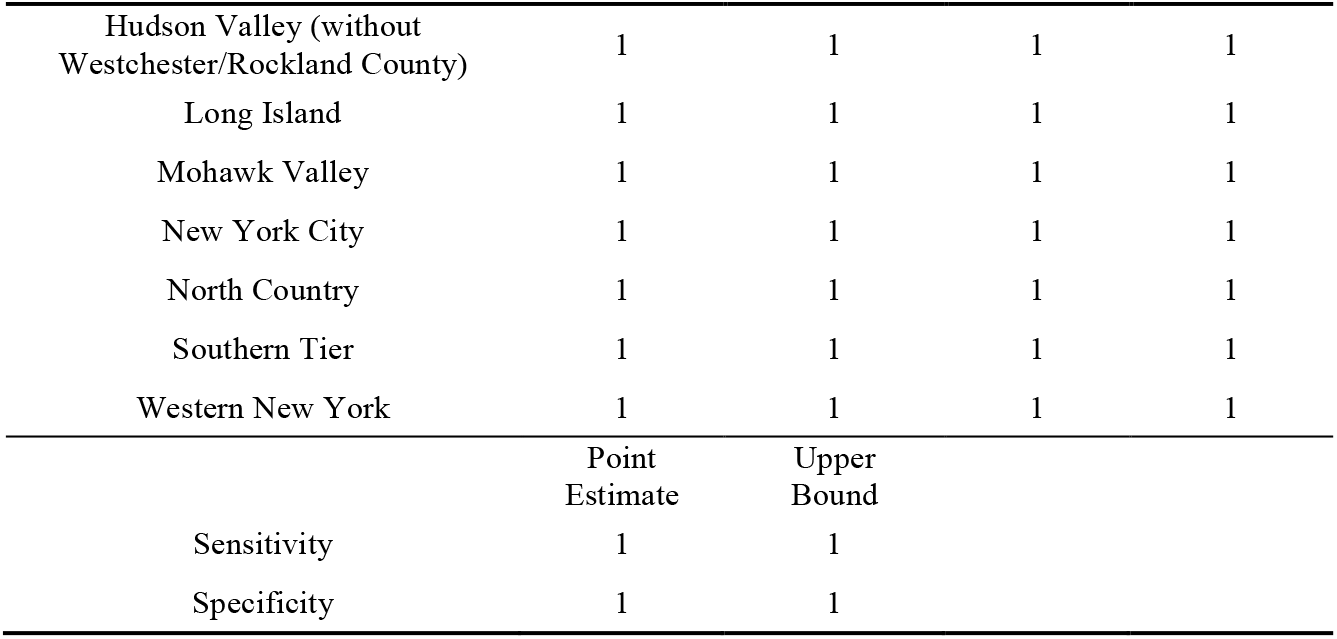
Convergence diagnostics for the uniform prior prevalence model. The table contains point estimates and upper bound of the 95% credible interval for the Gelman-Rubin Diagnostic or potential scale reduction factor. Values of 1 suggest convergence.

**Table 4:**
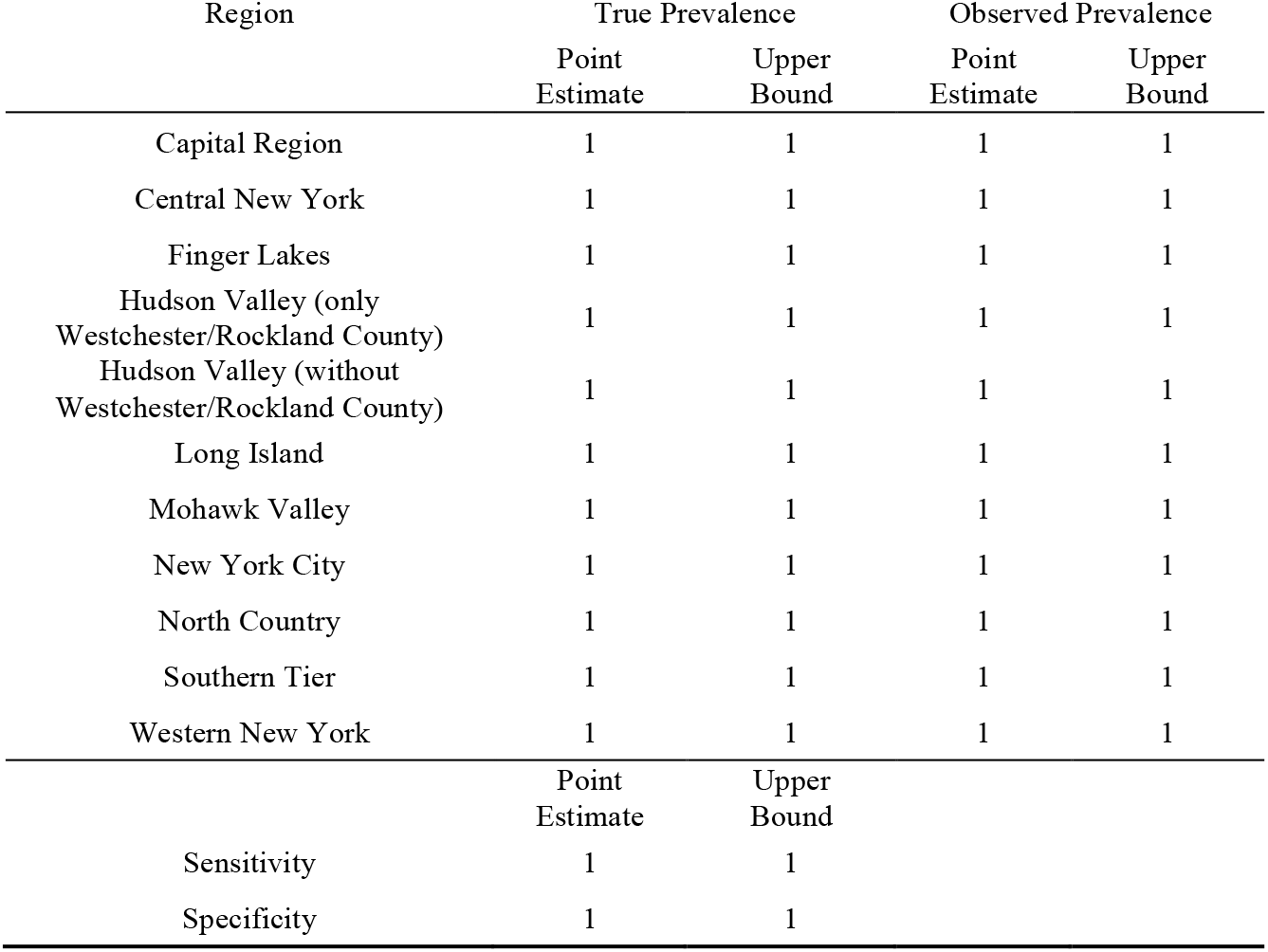
Convergence diagnostics for the informative prior prevalence model. The table contains point estimates and upper bound of the 95% credible interval for the Gelman-Rubin Diagnostic or potential scale reduction factor. Values of 1 suggest convergence.

**Fig 1.**
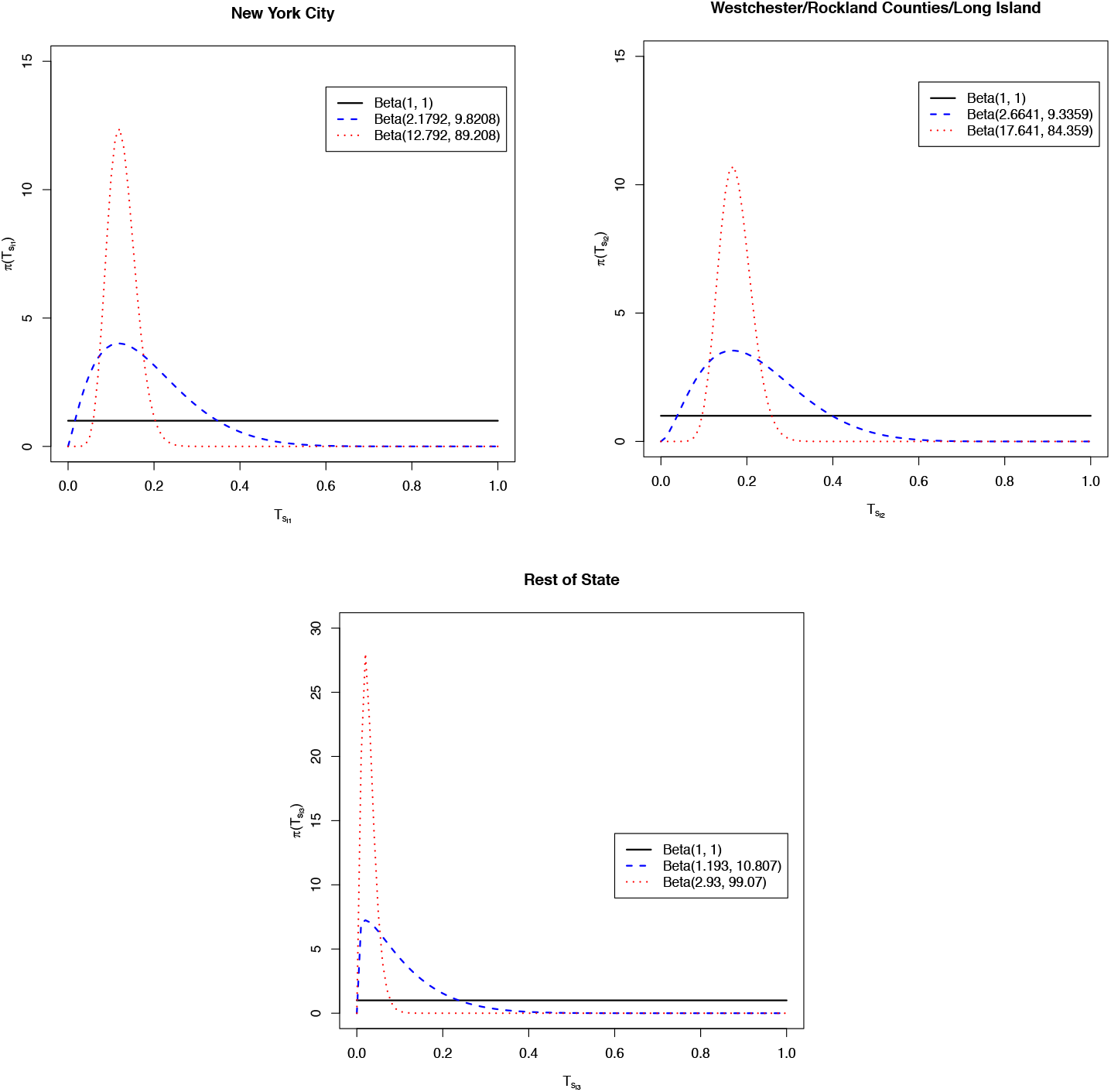
Plots of the prior distributions on true prevalence, *Ts*_*ij*_, by super-region. Uniform priors are in solid black, primary model (weakly informative) priors in dashed blue, and informative priors in dotted red. Specific prior details on provided in each plot’s legend.

**Fig 2.**
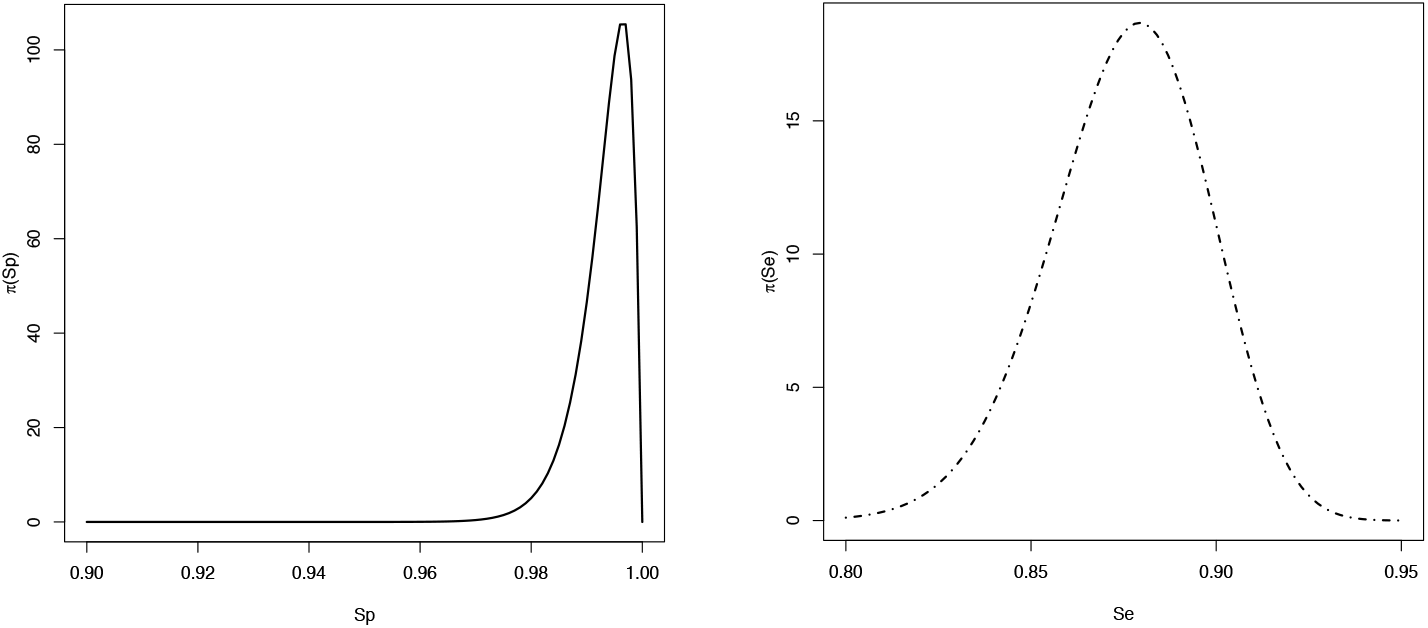
Plots of the prior distributions on specificity, *Sp* (left panel), and sensitivity *Se* (right panel).

### Primary Model (Weakly Informative Prior)

**Fig 3.**
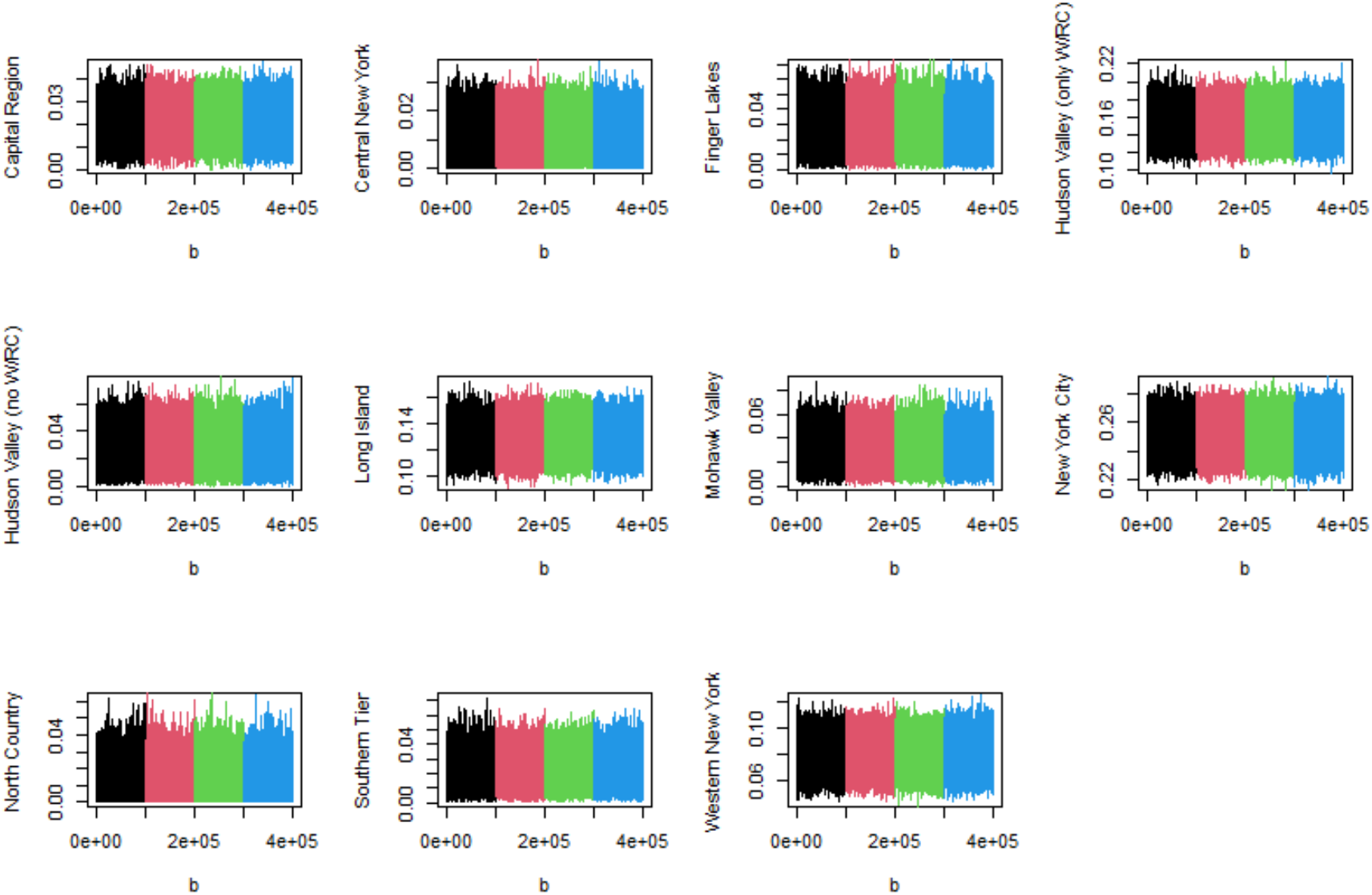
Traceplots of the posterior true prevalence, by region, for the primary model.

**Fig 4.**
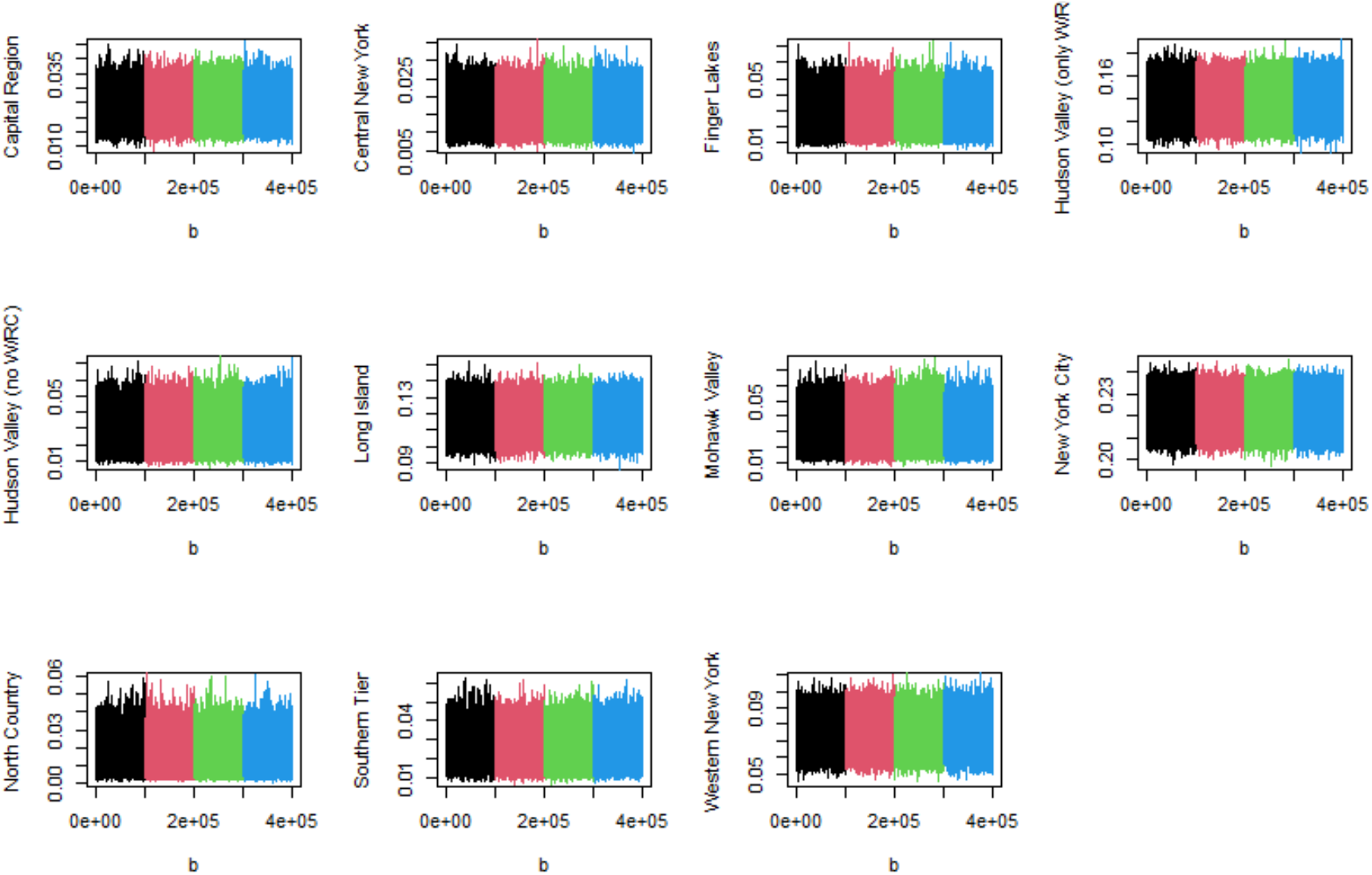
Traceplots of the posterior observed prevalence, by region, for the primary model.

**Fig 5.**
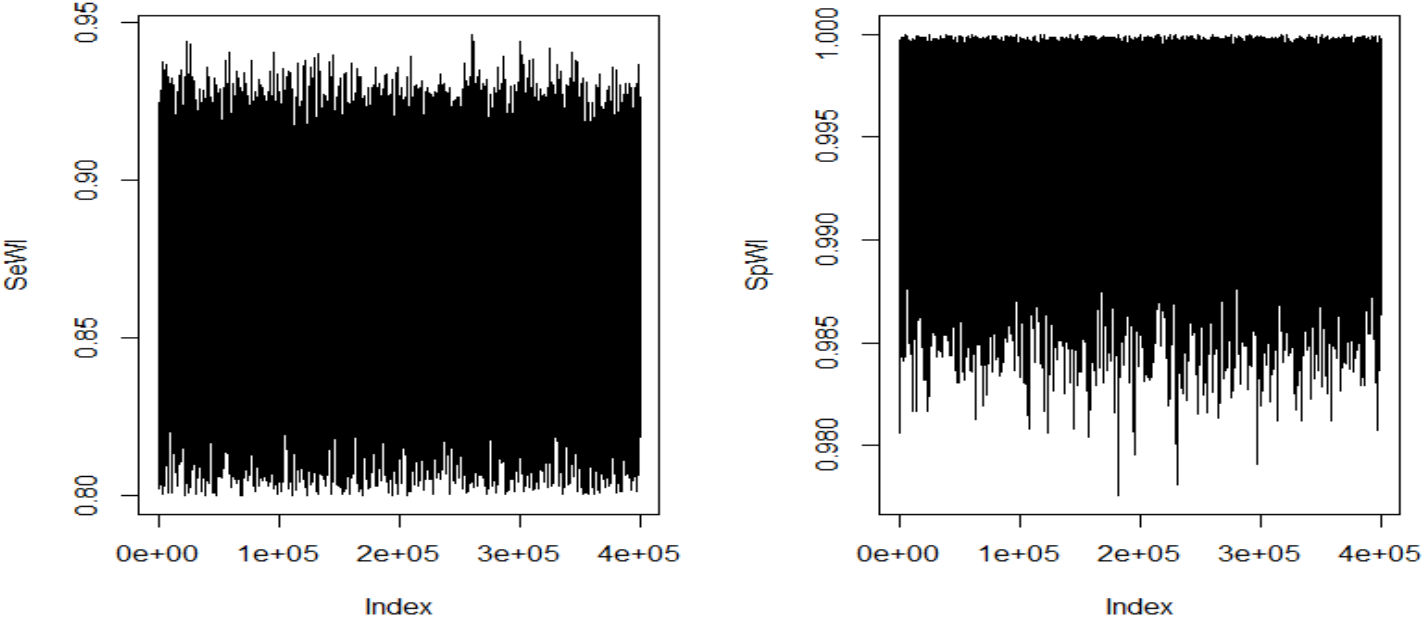
Traceplots of the posterior sensitivity and specificity for the primary model.

### Uniform Prior Prevalence Model

**Fig 6.**
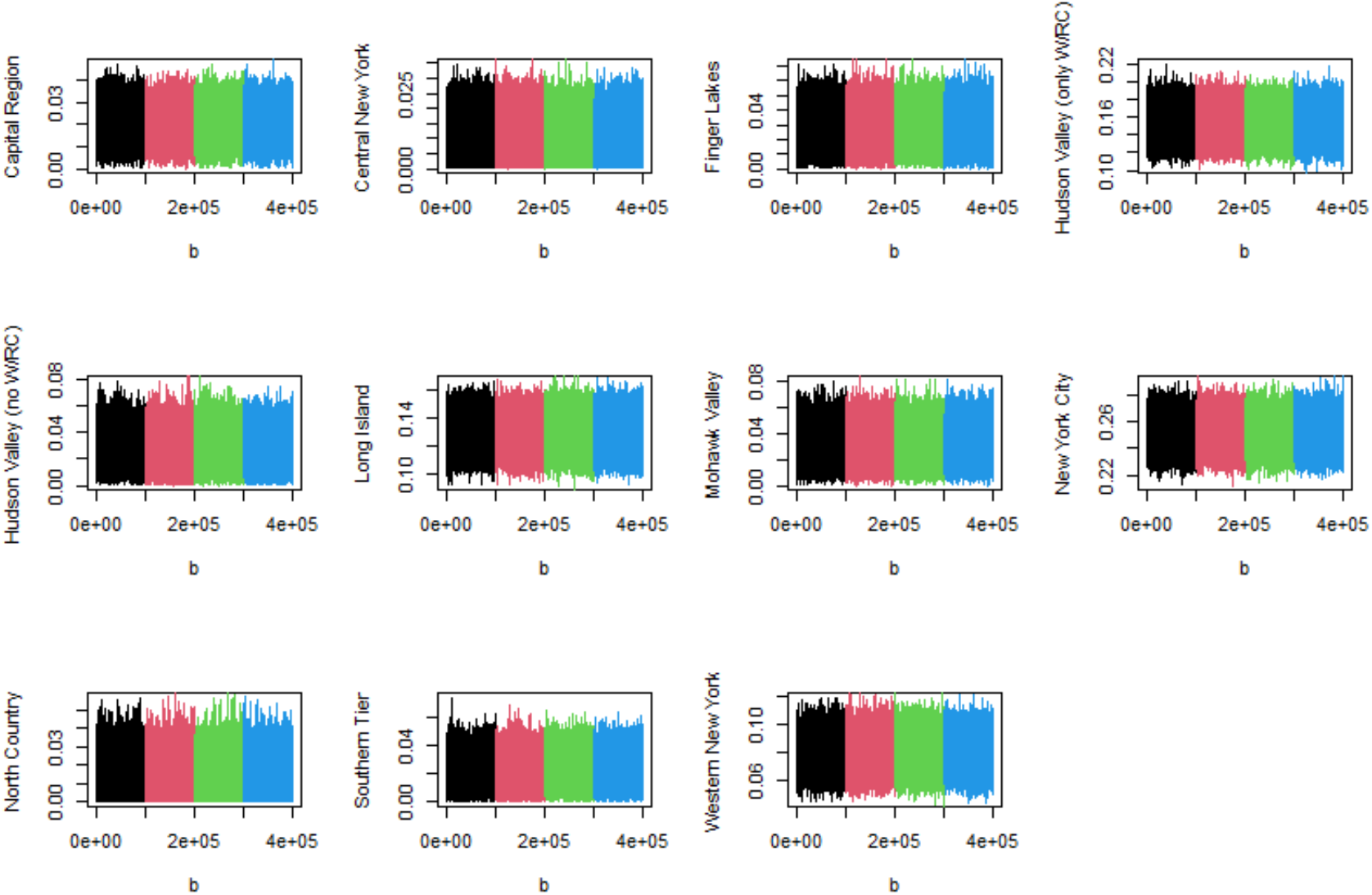
Traceplots of the posterior true prevalence, by region, for the uniform prior prevalence model.

**Fig 7.**
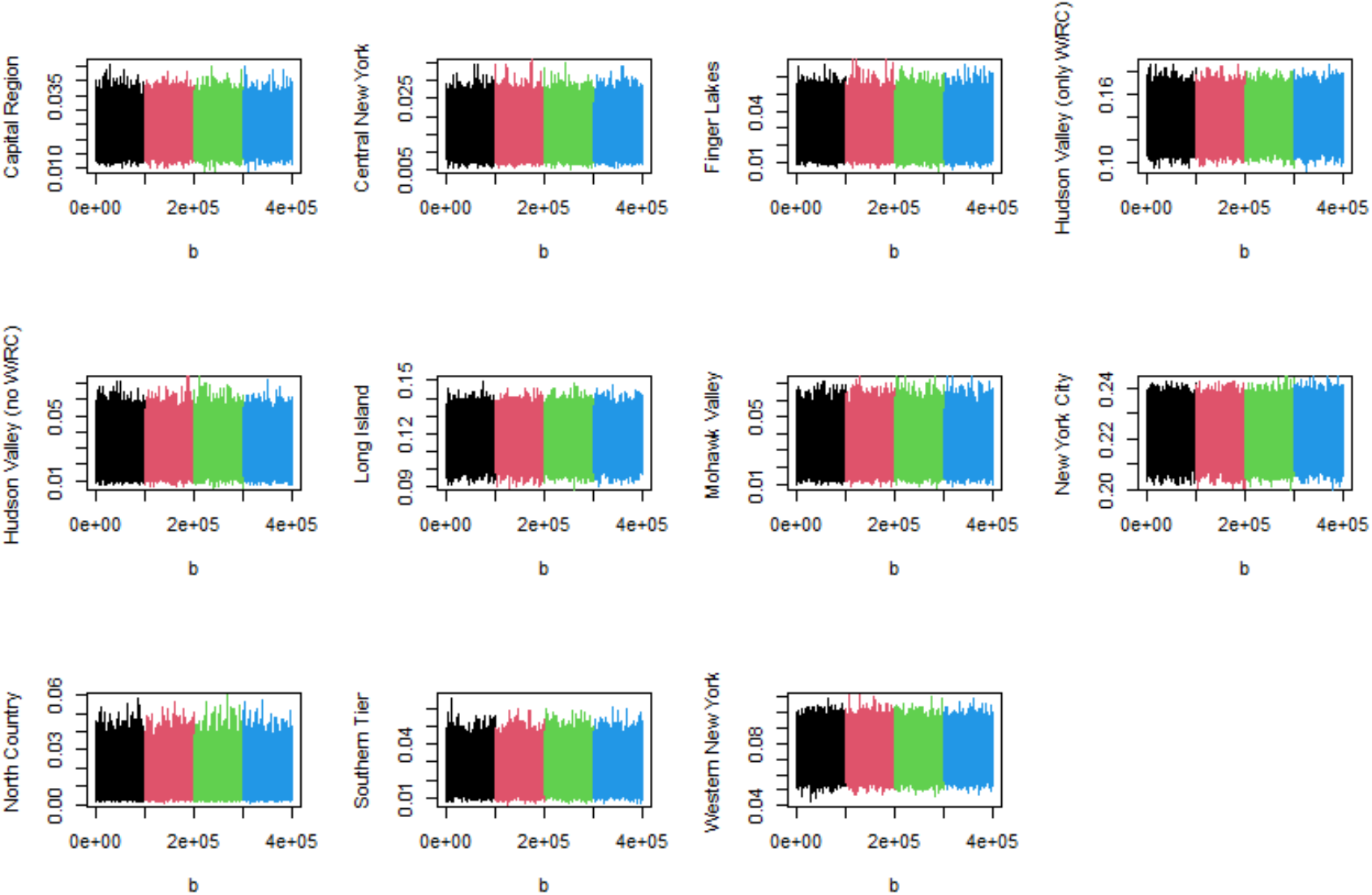
Traceplots of the posterior observed prevalence, by region, for the uniform prior prevalence model.

**Fig 8.**
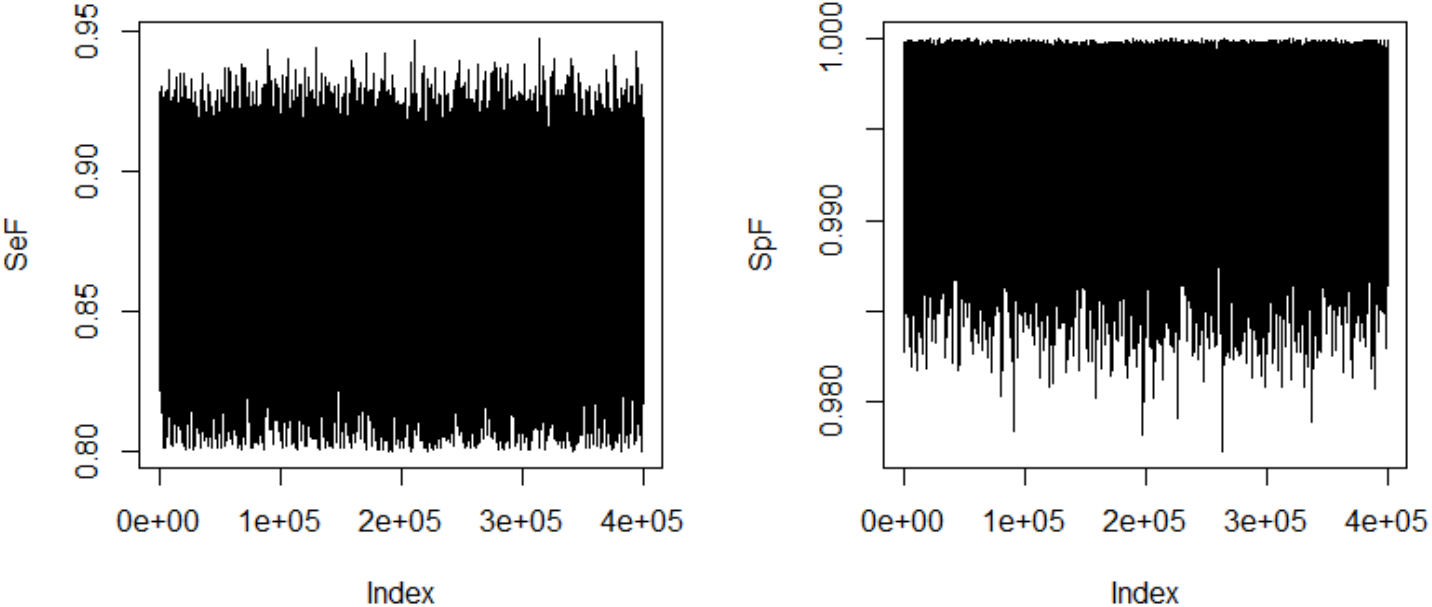
Traceplots of the posterior sensitivity and specificity for the uniform prior prevalence model.

### Informative Prior Prevalence Model

**Fig 9.**
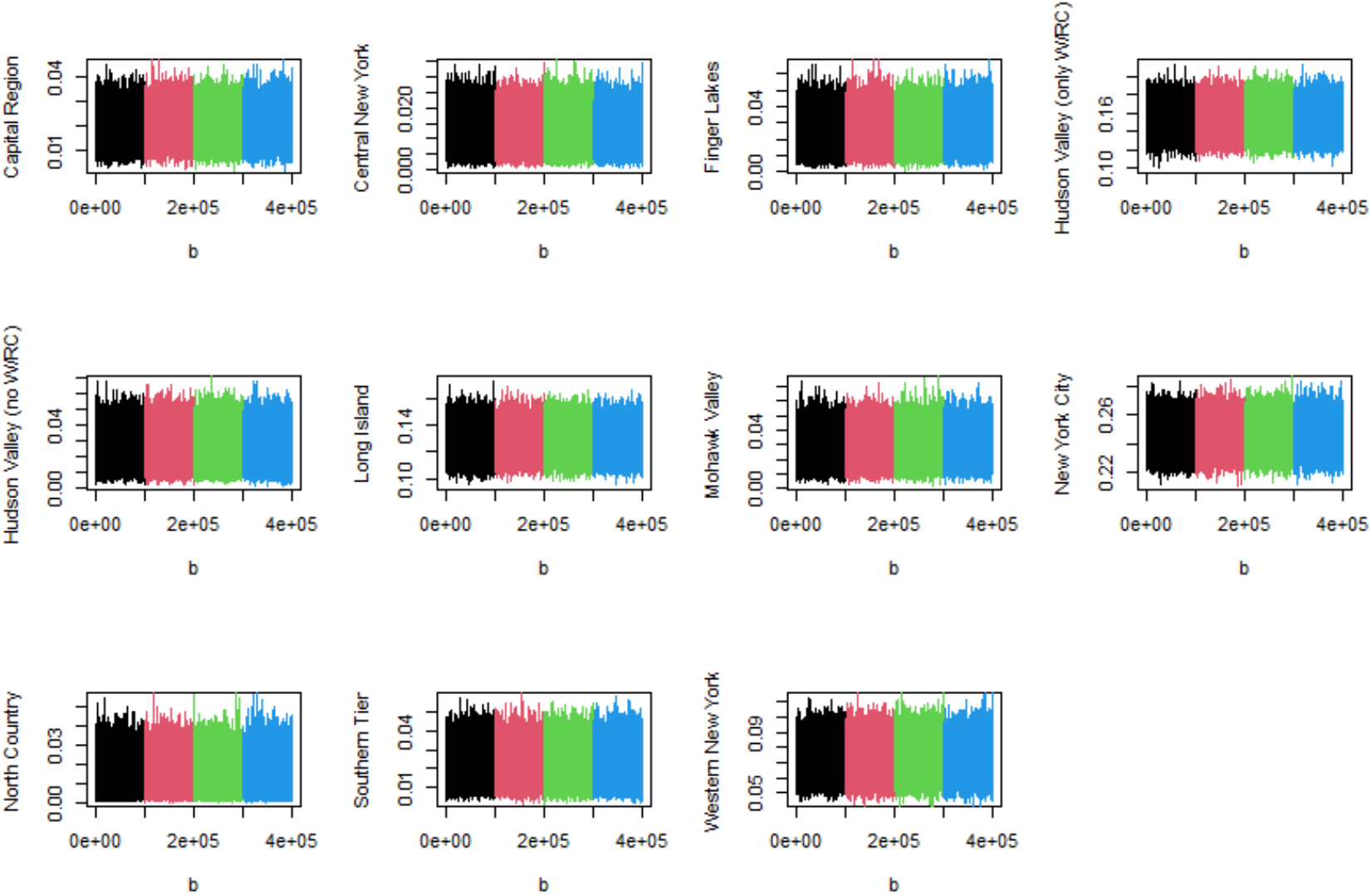
Traceplots of the posterior true prevalence, by region, for the informative prior prevalence model.

**Fig 10.**
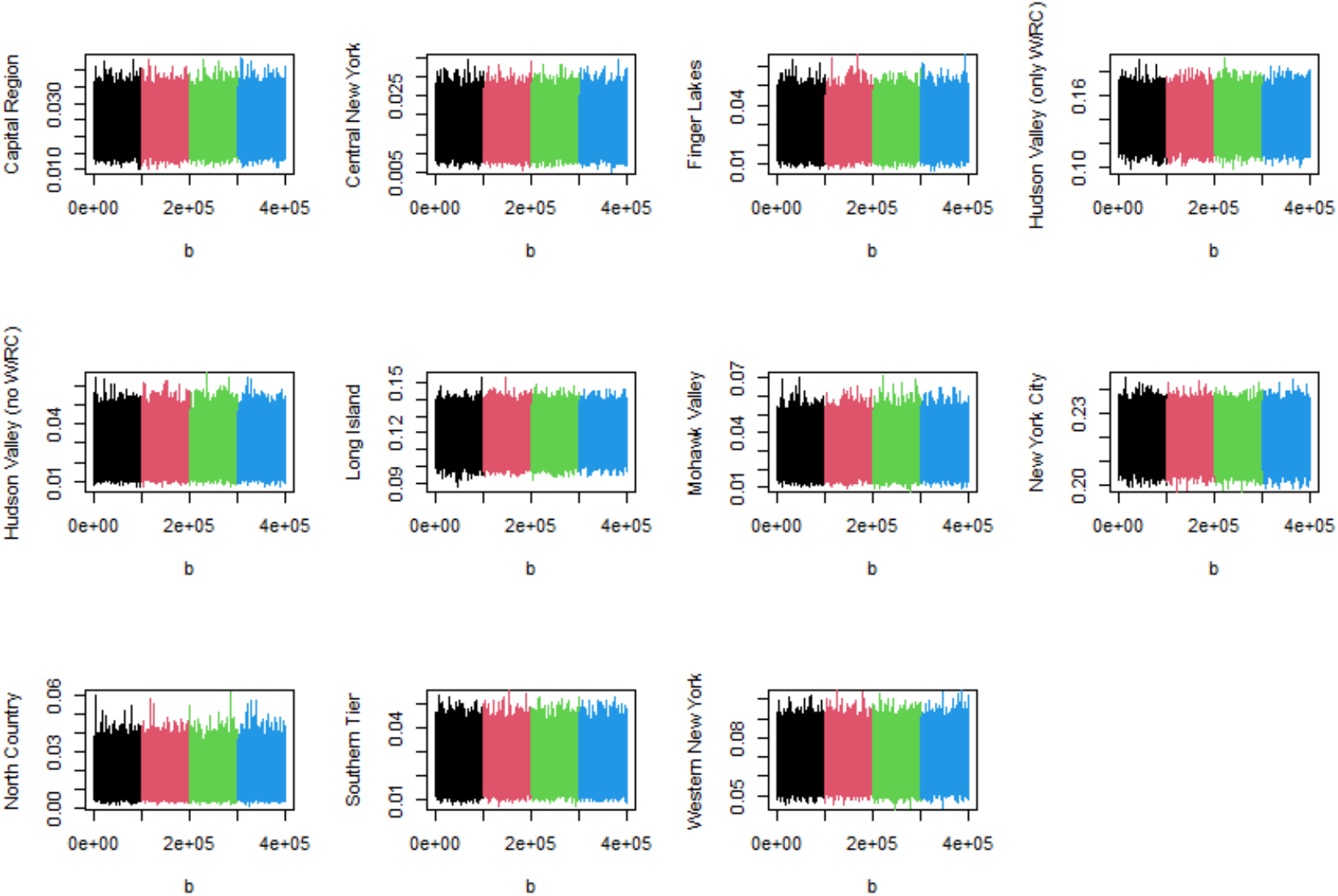
Traceplots of the posterior observed prevalence, by region, for the informative prior prevalence model.

**Fig 11.**
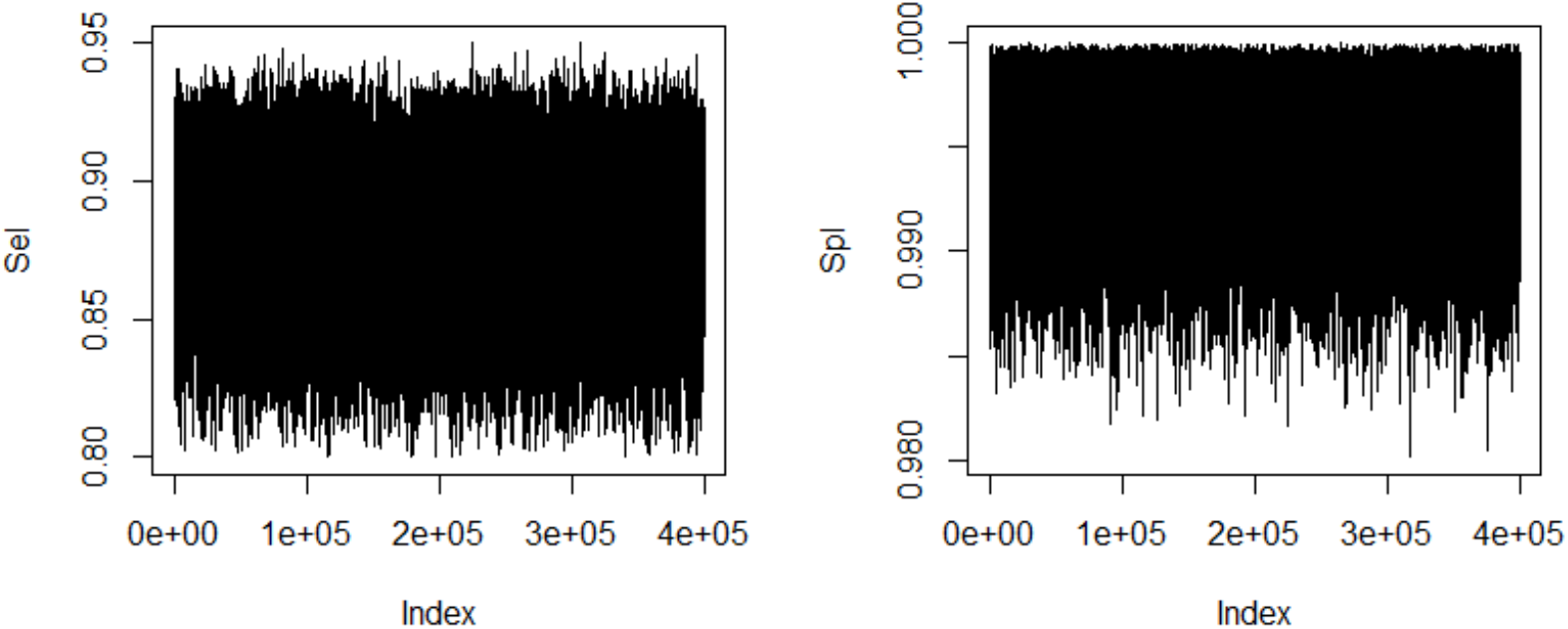
Traceplots of the posterior sensitivity and specificity for the informative prior prevalence model.

## Notes

### Competing Interest Statement

The authors have declared no competing interest.

### Funding Statement

This methods study had no external funding.

### Author Declarations

This study did not require IRB approval because it only used aggregate data already reported; the analysis did not use any individual-level data.

